# Natural history of patients with autosomal dominant *WFS1* pathogenic variants associated with sensorineural hearing loss and optic atrophy

**DOI:** 10.1101/2025.02.23.25322342

**Authors:** Jessica P. Roberts, Abby F. Tang, Daniela Hernandez, Brianna Carman, Liam Oiknine, Cris Brown, Stacy Hurst, Fumihiko Urano

**Affiliations:** Department of Medicine, Division of Endocrinology, Metabolism, and Lipid Research, Washington University School of Medicine, 660 South Euclid Avenue, St. Louis, MO 63110, USA; Department of Pathology and Immunology, Washington University School of Medicine, 660 South Euclid Avenue, St. Louis, MO 63110, USA; Georgetown University School of Medicine, Washington, DC, USA

**Author notes:** Correspondence information: Fumihiko Urano, MD, PhD: 660 South Euclid Ave, MSC 8127-0021-09, St. Louis, Missouri 63110, USA. Denotes corresponding author (Fumihiko Urano).

## Abstract

**Objective:** Autosomal dominant pathogenic variants in the WFS1 gene can cause a broad spectrum of WFS1-related disorders. These disorders present with a range of phenotypic manifestations, including isolated low-frequency sensorineural hearing loss, optic nerve atrophy accompanied by low- to mid-frequency sensorineural hearing loss, isolated diabetes mellitus, and early-onset cataracts. In general, WFS1-related disorders represent a milder spectrum of conditions linked to pathogenic WFS1 variants, except for Hattersley-Urano syndrome, which is characterized by early-onset diabetes mellitus, optic nerve atrophy, cataracts, hypotonia, intellectual disability, and developmental delay. By contrast, autosomal recessive WFS1 variants result in Wolfram Syndrome type 1, a rare neurodegenerative disorder characterized by early-onset diabetes mellitus, optic nerve atrophy, arginine vasopressin deficiency, hearing loss, and cerebellar and brainstem atrophy. Although WFS1-related disorders have been increasingly recognized, additional data are needed to understand their clinical progression and long-term outcomes. Our study aims to expand knowledge on the severity and progression of WFS1-related disorders by reviewing clinical data from patients with autosomal dominant pathogenic WFS1 variants.

**Approach:** We obtained clinical data from the Washington University International Registry and Clinical Study for Wolfram Syndrome and related disorders and the Endoplasmic Reticulum Disease Patient Registry and Biorepository. We included participants with autosomal dominant WFS1 pathogenic variants who were diagnosed with optic nerve atrophy and sensorineural hearing loss. Eleven participants with autosomal dominant WFS1 variants meeting these criteria were identified.

**Results:** The 11 cases included five distinct autosomal dominant WFS1 variants: c.923C>G (p.Ser308Cys), c.2051C>T (p.Ala684Val), c.2389G>T (p.Asp797Tyr), c.2456A>C (p.Gln819Pro), and c.2590G>A (p.Glu864Lys). Among these, the p.Gln819Pro variant has not been previously reported in the literature. The median age of optic atrophy diagnosis was 10 years (quartiles: 6.0 and 19.0 years). Visual acuity did not significantly differ between the left (OS) and right (OD) eyes (p = 0.8901). The least square best-corrected visual acuity (BCVA) mean for the right eye was 0.2114 ± 0.01903 and for the left eye, 0.2153 ± 0.01903. Age was not significantly related to best eye BCVA (p = 0.9196), with an estimated change of -0.0002 (95% CI [-0.003, 0.003]) per year. Patient age was also not correlated with binocular BCVA (p = 0.5994), with an estimated change of 0.00075 (95% CI [-0.0021, 0.0036]) per year. Mean retinal nerve fiber layer (RNFL) thickness was not significantly related to age (p = 0.1604), with an estimated annual change of 0.1486 (95% CI [-0.659, 0.363]). However, removing an influential outlier resulted in a significant relationship between RNFL thickness and age (p = 0.0160), with an estimated change of 0.2114 (95% CI [0.045, 0.377]) per year. Hearing loss diagnoses occurred at a median age of 2.0 years (quartiles: 1.5 and 2.0 years). All participants used hearing aids (11/11); six (6/11) had cochlear implants, while three (3/11) used external hearing aids. The median time between hearing loss diagnosis and hearing aid use was 4.0 years (quartiles: 2.5 and 8.0 years).

**Conclusion:** This study contributes to the growing understanding of WFS1-related disorders caused by autosomal dominant WFS1 variants. In particular, it highlights two clinical phenotypes of a novel WFS1 variant and provides valuable insights into the progression of optic nerve atrophy and hearing loss management.

## Introduction

The WFS1 gene was originally identified in 1998 as the causative gene for Wolfram syndrome, a rare autosomal recessive genetic disorder characterized by antibody-negative early-onset diabetes mellitus, arginine vasopressin deficiency, optic nerve atrophy, and sensorineural hearing loss, along with various other neurological and psychological features (1–4). Over the past decade, human disorders associated with dominant variants of the WFS1 gene have received increasing attention. These conditions, often referred to as Wolfram-like diseases or syndromes, have become more diverse and are generally clinically less severe compared to Wolfram syndrome. As a result, the broader term “WFS1-related disorders” is now commonly used. The clinical features associated with WFS1-related disorders can vary widely. These features include isolated low-frequency sensorineural hearing loss, optic nerve atrophy accompanied by low to mid-frequency sensorineural hearing loss, isolated diabetes mellitus, and isolated early-onset cataracts. While most autosomal dominant WFS1-related disorders are generally milder than Wolfram syndrome, there is a severe disorder known as Hattersley-Urano syndrome. This syndrome, caused by autosomal dominant WFS1 variants, is characterized by neonatal diabetes, congenital sensorineural deafness, congenital cataracts, hypotonia, developmental delay, and intellectual disability (5).

Although the natural history of Wolfram syndrome has been extensively studied, the natural history of WFS1-related disorders has received limited research attention. One specific constellation of clinical features in WFS1-related disorders includes optic nerve atrophy and sensorineural hearing loss, which represents the less severe end of the spectrum of disorders caused by WFS1 variants (6–9). In this study, we present the clinical features of 11 patients with autosomal dominant WFS1 variants. Specifically, we focus on metrics of patients’ visual and hearing health. Thus, we have included visual acuity and retinal nerve fiber layer (RNFL) thickness measures, and hearing aid use. These findings will add to a small but growing body of literature on the visual and hearing impairments associated with WFS1-related disorder (6, 7, 10, 11).

## Materials and Methods

### Patient clinical information and genetic analysis

Patient clinical information was obtained from the Washington University International Registry and Clinical Study for Wolfram Syndrome and the Endoplasmic Reticulum Disease Patient Registry and Biorepository. The minimum diagnostic inclusion criteria: (1) Genetic analysis confirmation of an autosomal dominant variant in the *WFS1* gene. (2) Formal diagnosis of optic atrophy and sensorineural hearing loss. Subjects, or their parent or legal guardian, provided signed written, informed consent for participation in the study and release of personal health information prior to their inclusion in this study. This investigation was approved by the Human Research Protection Office at Washington University School of Medicine in St. Louis, MO (IRB IDs #201107067 and #201807044).

### Clinical assessment and data collection

Clinical information on the 11 participants was gathered from the Washington University International Registry and Clinical Study for Wolfram Syndrome, Endoplasmic Reticulum Disease Patient Registry and Biorepository, and patients’ medical records. Collected records include age, sex, *WFS1* variant, age at optic atrophy diagnosis, age at hearing loss diagnosis, hearing aid use, age at hearing aid installation, other vision conditions, best corrected distance visual acuity (BCVA), and RNFL thickness measurements.

### Statistical Analysis

Proc Mixed of SAS software was used for Windows (V9.4) to implement a repeated measures model to analyze differences in visual acuity and RNFL thickness between patients’ eyes and account for repeated measurements from the same patient. This software was also used to test for a significant relationship between age and best eye BCVA, BCVA OU, and mean RNFL thickness adjusting for multiple observations per participant at variable ages.

## Results

### Participant Demographics and Clinical Features

We identified 11 patients in the Washington University International Registry and Clinical Study for Wolfram Syndrome and the Endoplasmic Reticulum Disease Patient Registry and Biorepository with autosomal dominant variants in the *WFS1* gene that have been diagnosed with optic nerve atrophy and sensorineural hearing loss. A summary of the demographics and clinical features of these patients is provided in **Table 1**.

**Table 1.** Summary of autosomal dominant *WFS1* variant cases.

| Case | Sex | Age (years) | cDNA variant | Protein variant | Age at optic atrophy diagnosis (years) | Age at hearing loss diagnosis (years) | Hearing aid use | Age at Hearing Aid Installation (Right \| Left) | Other Vision Conditions |
| --- | --- | --- | --- | --- | --- | --- | --- | --- | --- |
| 1 | M | 15 - 19 y | c.2051C>T | p.Ala684Val | 5 - 9 y | 0 - 4 y | Y - cochlear implants | 0 - 4 y \| 0 - 4 y | Myopia |
| 2 | M | 25 - 29 y | c.2051C>T | p.Ala684Val | 15 - 19 y | 0 - 4 y | Y | NA | Myopia, astigmatism, reduced color perception |
| 3 | M | 20 - 24 y | c.2389G>T | p.Asp797Tyr | 10 - 14 y | 0 - 4 y | Y - cochlear implants | 0 - 4 y \| 5 - 9 y |  |
| 4 | M | 50 - 54 y | c.923C>G | p.Ser308Cys | 40 - 44 y | 0 - 4 y | Y - external aids | 35 - 39 y \| 35 - 39 y |  |
| 5 | F | 15 - 19 y | c.2051C>T | p.Ala684Val | 15 - 19 y | 0 - 4 y | Y - cochlear implants | 10 - 14 y \| 10 - 14 y | Color blindness |
| 6 | F | 40 - 44 y | c.2590 G>A | p.Glu864Lys | 10 - 14 y | 0 - 4 y | Y | NA | Color vision deficiency |
| 7 | F | 5 - 9 y | c.2051C>T | p.Ala684Val | 5 - 9 y | 0 - 4 y | Y - cochlear implants | 0 - 4 y \| 5 - 9 y |  |
| 8 | M | 5 - 9 y | c.2051C>T | p.Ala684Val | 0 - 4 y | 0 - 4 y | Y - cochlear implants | 0 - 4 y \| 0 - 4 y |  |
| 9 | M | 5 - 9 y | c.2051C>T | p.Ala684Val | 0 - 4 y | 0 - 4 y | Y - cochlear implants | 0 - 4 y \| 0 - 4 y |  |
| 10 | F | 40 - 44 y | c.2456 A>C | p.Gln819Pro | 40 - 44 y | 5 - 9 y | Y - external aids | NA | Myopia, elevated intraocular pressure, color blindness, peripheral visual field constriction |
| 11 | F | 5 - 9 y | c.2456 A>C | p.Gln819Pro | 5 - 9 y | 0 - 4 y | Y - external aids | 0 - 4 y \| 0 - 4 y | Myopia, astigmatism |

The median age of patients in this study is 19.0 years (lower and upper quartiles: 9.0 and 43.0 years, respectively). The median age at optic atrophy diagnosis is 10 years (6.0 and 19.0 years). Four patients have myopia (4/11), four patients have color vision deficiencies (4/11), two patients have astigmatism (2/11), and one patient (1/11) has elevated intraocular pressure and peripheral visual field constriction.

For sensorineural hearing loss diagnosis, the median age is 2.0 years (1.5 years and 2.0 years). All patients (11/11) in this study use hearing devices to aid their sensorineural hearing loss. Six patients (6/11) use cochlear implants, three (3/11) use external hearing aids, while the form of hearing aid used by the remaining two patients (2/11) is unknown. Patients began using hearing aids at a median age of 4.0 years (2.5 years and 8.0 years). Of the eight patients with known hearing aid installation dates, the median time from hearing loss diagnosis to first hearing aid installation is 2.38 years (2.0 years and 6.4 years).

### WFS1 Variant Genotypes and Predicted Domain Locations

The five autosomal dominant *WFS1* variants included in this study are listed in **Table 2**. Six (6/11) patients possess the c.2051C>T (p.Ala684Val) *WFS1* variant, making it the most prevalent in the cohort. Patients 10 and 11 (2/11) share a novel c.2456A>C (p.Gln819Pro) variant, which Patient 11 maternally inherited from Patient 10. Furthermore, one patient (1/11) has a c.923C>G (p.Ser308Cys) variant, one (1/11) patient has a c.2389G>T (p.Asp797Tyr) variant, and one (1/11) patient has a c.2590G>A (p.Glu864Lys) variant. All included variants are missense. Four of the altered amino acids have a predicted location in the endoplasmic reticulum (ER) luminal domain of the *WFS1* protein, whereas the p.Ser308Cys variant is likely positioned within the cytosolic *WFS1* domain (12).

**Table 2.** *WFS1* variants in study cohort The five heterozygous *WFS1* variants included in this study are shown in Table 2. The p.Ala684Val variant is the most prevalent in this cohort as six patients (6/11) have this genotype. Patients 10 and 11 share a novel p.Gln819Pro variant (2/11), which patient 11 maternally inherited from patient 10. Furthermore, one patient has a p.Ser308Cys variant, one patient has a p.Asp797Tyr, and one patient has a p.Glu864Lys variant. ER = endoplasmic reticulum.

| cDNA | Protein | Domain | Variant type | Prior reports |
| --- | --- | --- | --- | --- |
| c.923C>G | p.Ser308Cys | cytosolic | missense | Yes |
| c.2051C>T | p.Ala684Val | ER lumen | missense | Yes |
| c.2389G>T | p.Asp797Tyr | ER lumen | missense | Yes |
| c.2456 A>C | p.Gln819Pro | ER lumen | missense | No |
| c.2590 G>A | p.Glu864Lys | ER lumen | missense | Yes |

### Visual Acuity

To assess the progression of patients’ optic atrophy, we identified best-corrected distance logMAR visual acuity (BCVA) measures recorded during nine (9/11) patients’ optometry and ophthalmology visits. There was no overall difference in visual acuity between patients’ left (OS) and right (OD) eyes (p = 0.8901). The least square BCVA mean for the right eye (OD) was 0.2114 ± 0.01903, while for the left eye, it was 0.2153 ± 0.01903. Given that there was no significant discrepancy in relative eye visual acuity, we used the BCVA of both eyes (OU) and best eye for further analyses. In **Figure 1a**, BCVA of best eye (based on first visit) is illustrated as a function of age. Age was not significantly related to best eye BCVA (p=0.9196). The estimated change in best eye BCVA with age was -0.0002 (95% CI [-0.003, 0.003]). Furthermore, we investigated trends in BCVA OU of the same nine (9/11) patients at various ages, which is illustrated in **Figure 1b**. Patient age was not significantly correlated with BCVA OU (p = 0.5994). The estimated change in BCVA OU with age was 0.00075 (95% CI [-0.0021, 0.0036]). Of note, Patient 1 has myopia, Patient 2 has myopia and astigmatism, Patient 10 has myopia, elevated intraocular pressure, and peripheral visual field constriction, and Patient 11 has myopia and astigmatism. These additional visual conditions may influence the BCVA of these individuals.

**Figure 1a.**
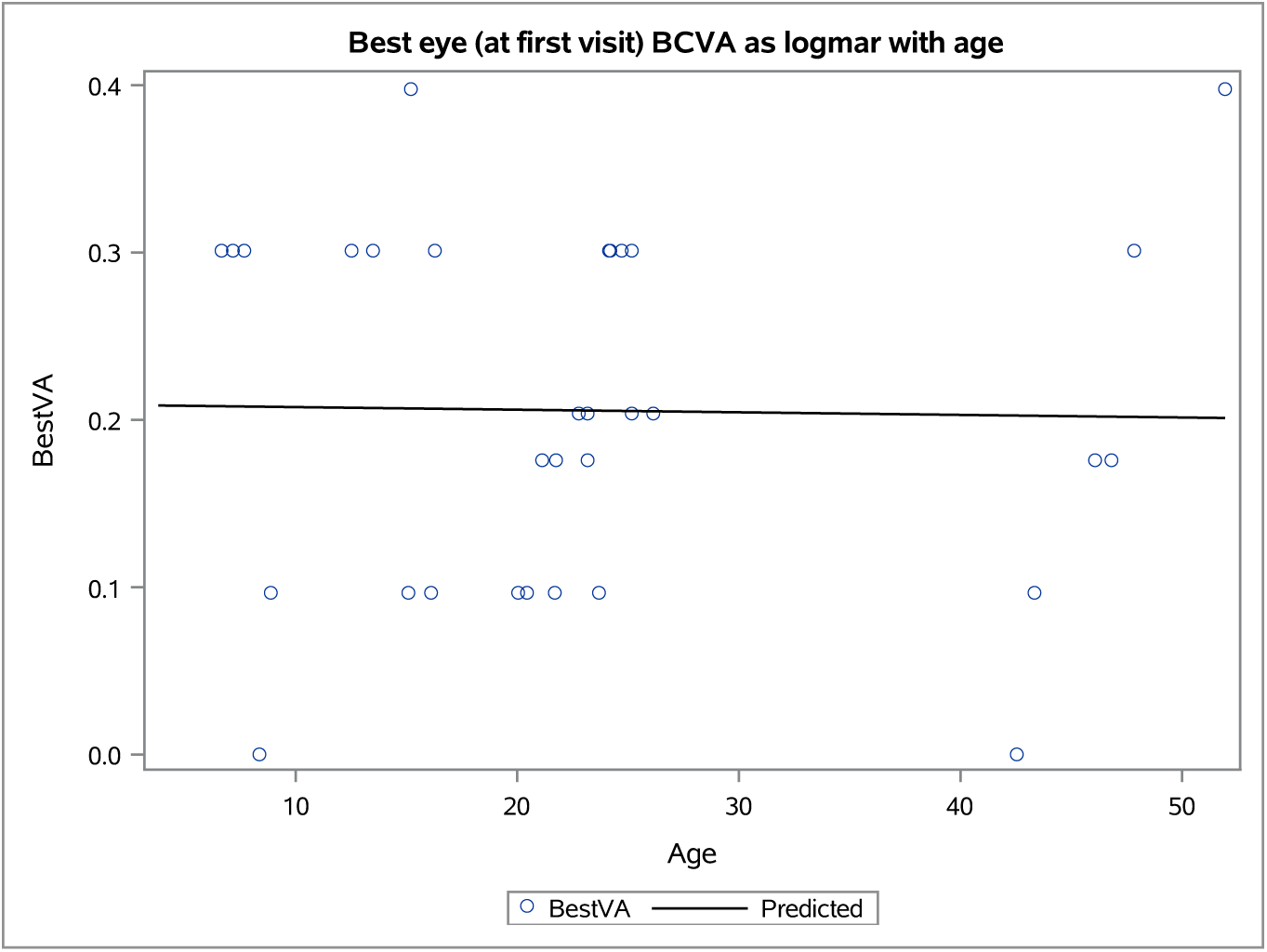
Best-corrected visual acuity of best eye versus patient age at first visit Best-corrected visual acuity (BCVA) of best eye at first visit (measured as logMAR) for nine patients versus age (measured in years). Age and best eye BCVA are not significantly correlated (p=0.9196). The solid line represents the slope estimate for age vs. BCVA, slope = - 0.0002 (95% CI [-0.003, 0.003]).

**Figure 1b.**
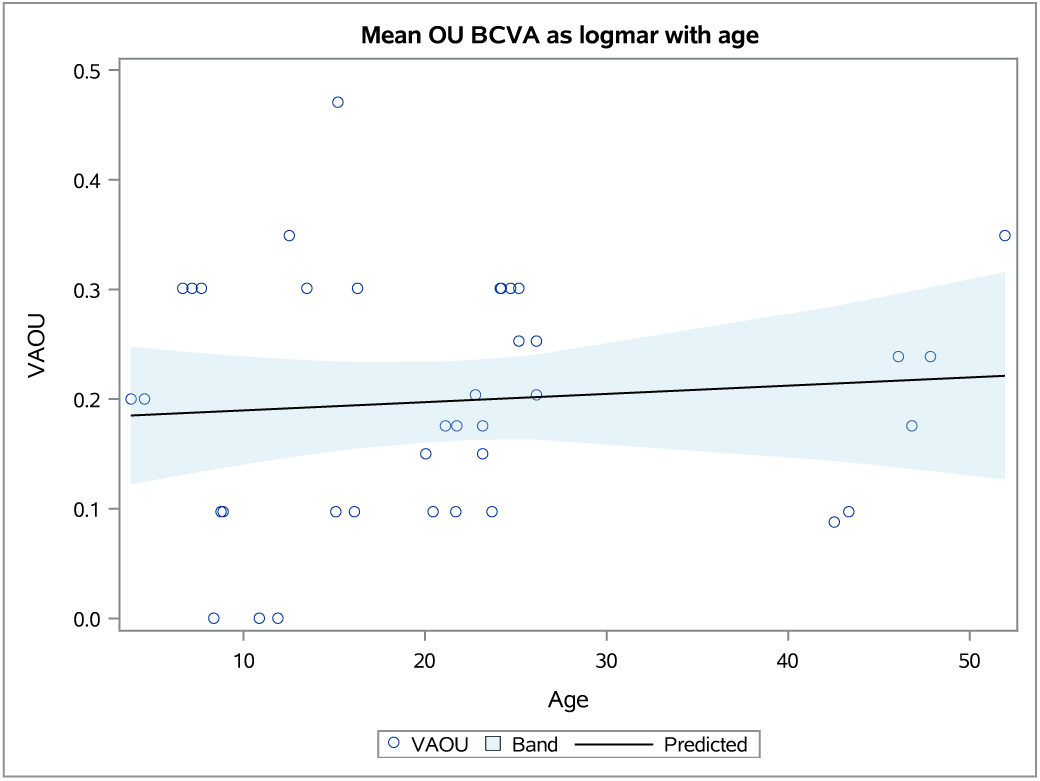
Visual acuity change with age Best-corrected binocular visual acuity (BCVA or VAOU) measured as logMAR of nine patients with age (years). Age was not significantly correlated with BCVA OU (p = 0.5994). The solid line represents the estimated slope for age versus VAOU, slope = 0.00075 (95% CI [- 0.0021, 0.0036]).

### Retinal Nerve Fiber Layer Thickness

In addition to visual acuity, retinal nerve fiber layer (RNFL) thickness can be used as a marker of optic atrophy in patients with autosomal dominant WFS1 related disorders based (6, 13, 14). Eight (8/11) patients in our cohort had recorded RNFL thickness measurements. **Figure 2a** depicts the mean RNFL thickness measurements adjusted for multiple observations per participant. Using all data, age was not significantly related to mean RNFL thickness (p=0.1604). The slope estimate is 0.1486 (95% CI [-0.659, 0.363]). Case 11 is an influential outlier in the RNFL analysis. Removing this outlier leads to a statistically significant relationship between mean RNFL and age (p=0.0160). The slope estimate is 0.2114 (95% CI [0.045, 0.377]) as shown in **Figure 2b**.

**Figure 2a.**
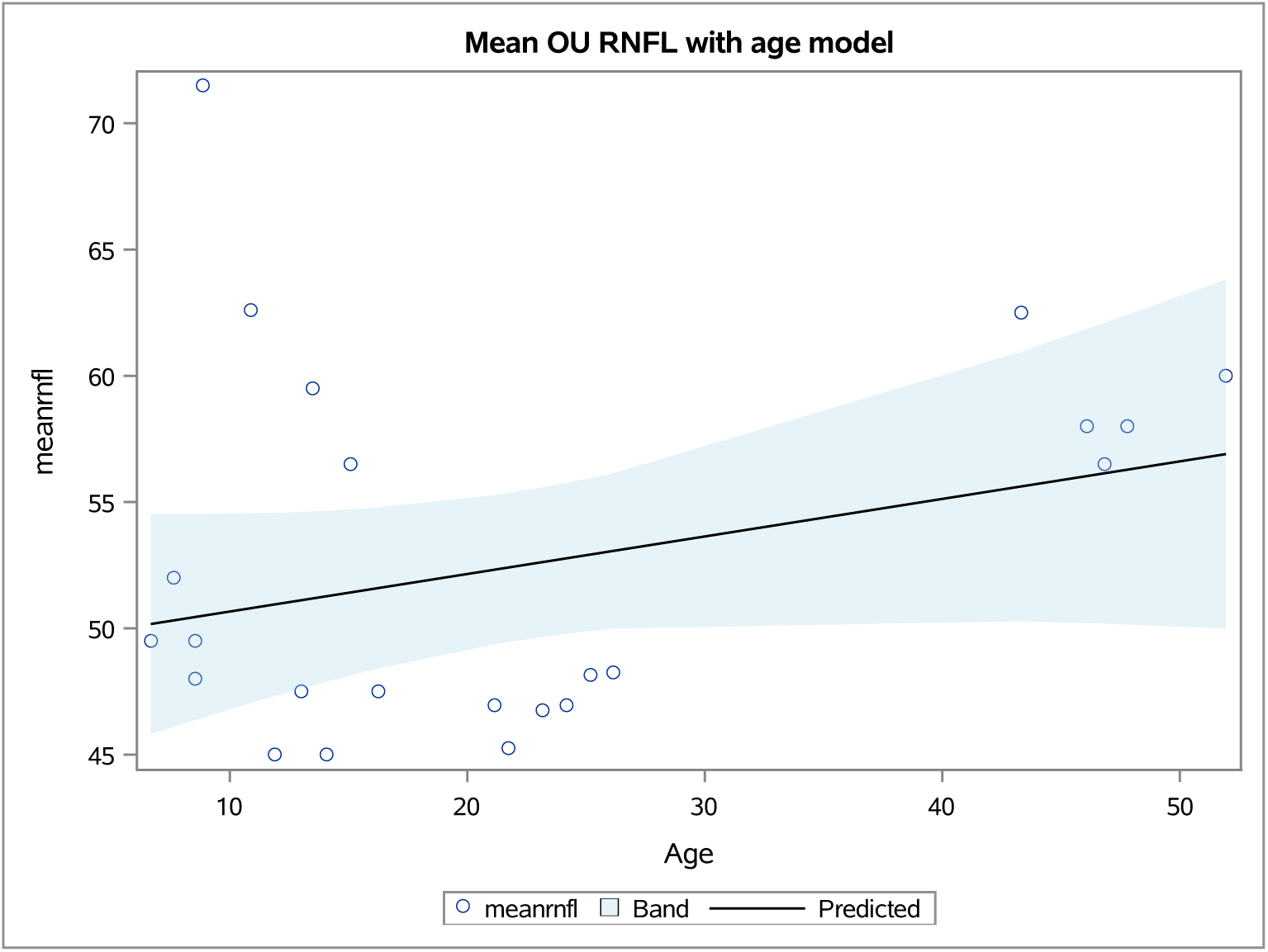
Mean RNFL Versus Age Mean Retinal Nerve Fiber Layer (RNFL) thickness (μm) calculated from right (OD) and left (OS) eye measurements for eight patients versus age (years). Age and RNFL thickness were not significantly correlated (p = 0.1604). The estimated slope = 0.1486 (95% CI [-0.659, 0.363]).

**Figure 2b.**
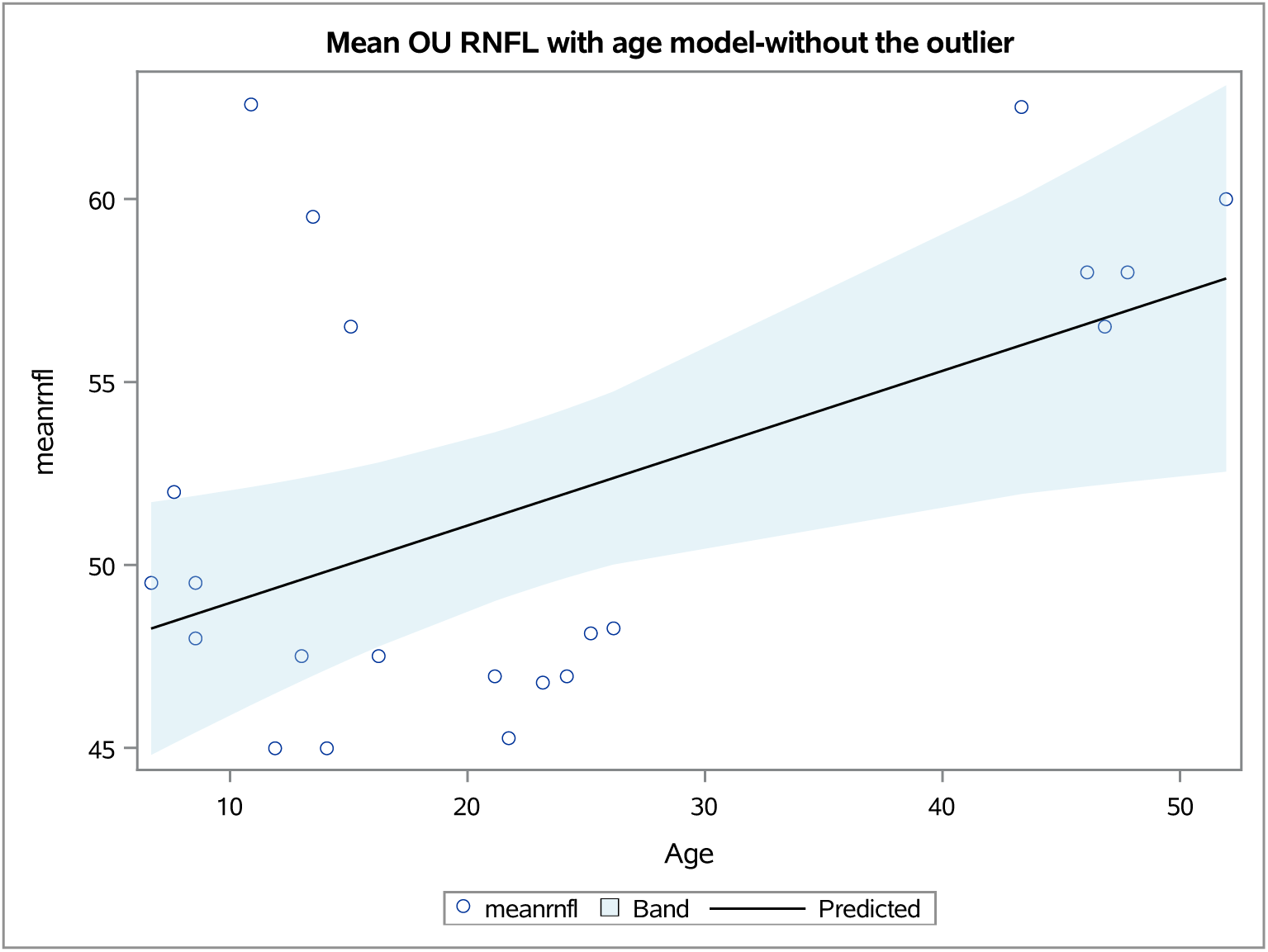
Mean RNFL vs. Patient Age (Outlier Excluded) Mean Retinal Nerve Fiber Layer (RNFL) thickness (μm) calculated from right (OD) and left (OS) eye measurements for seven patients versus age (years). With the removal of Case 11, age and mean RNFL thickness are significantly correlated (p = 0.0160). The estimated slope = 0.2114 (95% CI [0.045, 0.0377]).

## Discussion

Here, we have reported the clinical features of 11 patients with autosomal dominant WFS1 variants. The variants included are p.Ser308Cys, p.Ala684Val, p.Asp797Tyr, p.Gln819Pro, and p.Glu864Lys. The p.Gln819Pro variant has not been reported previously in the scientific literature.

Notably, all participants received sensorineural hearing loss diagnoses early in life, at a median age of 2.0 years and optic nerve atrophy diagnoses at a median age of 10.0 years. This timeline of sensorineural hearing loss diagnosis within the first few years of life and optic nerve atrophy diagnosis in the second decade of life is consistent with reported observations in the autosomal dominant *WFS1*-related disorder (Wolfram-like) patient population (6, 15, 16). This is in contrast to patients with autosomal recessive Wolfram syndrome patients, who initially experience high frequency hearing loss in their teens, which progresses to loss of lower frequencies with age (17). However, both autosomal recessive and autosomal dominant WFS1 variants lead to onset of optic atrophy in the early teen years (2, 3, 18).

Our analysis of BCVA in patients with autosomal dominant WFS1 variants suggests that BCVA remains relatively stable with age, indicating a slow progression of visual decline in WFS1-related disorders. Furthermore, BCVA might not be the ideal marker of optic atrophy progression due to the confounding effect of other visual conditions on this measurement. In our cohort, four patients had additional visual conditions that might alter visual acuity. Thus, we also examined the mean RNFL thicknesses.

The mean RNFL thickness for the eight patients in the cohort ranged from 45 to 71.5 μm, which is markedly below the mean RNFL thickness for healthy adults of a similar age range (6 to 51 years) (19). Although no significant correlation between mean RNFL thickness and age was observed when all eight patients were included, Patient 11 was identified as an outlier. When Patient 11’s mean RNFL thickness measurement was excluded, the estimated RNFL thickness increased by 0.2114 μm per year. Given that this finding contradicts the degenerative nature of WFS1-related disorder, this finding might be explained by the heterogeneity of autosomal dominant variants included in the cohort. Certain variants might lead to more extensive RNFL thinning. Notably, Patients 10 and 11, the patients with novel p.Gln819Pro variant have the third and first highest mean RNFL thicknesses in the cohort at 62.5 and 71.5 **μm**, respectively. Furthermore, these measurements were recorded when Patient 10 was between the ages of 40-44 years old, and Patient 11 was between the ages of 5-9 years old. Studies on autosomal recessive Wolfram Syndrome have also noted that RNFL thickness progression was correlated less with age and more with disease severity (20). Thus, it is possible that the p.Gln819Pro variant is associated with less severe manifestations of WFS1-related disorder. Another factor to consider for the estimated change in RNFL thickness with age is the RNFL thickness floor. This floor is the minimum thickness that optical coherence tomography (OCT) instruments can detect, which may lead to fluctuation of measurements taken for patients in this cohort with very low RNFL thicknesses (21). Overall, our analyses suggest that patients with WFS1-related disorders may initially present with reduced RNFL thickness. However, the progression of RNFL thinning is often minimal and appears to correlate more closely with disease severity than chronological age.

The clinical findings observed in our cohort are driven by underlying molecular mechanisms that underscore the essential role of WFS1 in maintaining cellular homeostasis and protecting against neurodegeneration. The WFS1 gene encodes wolframin (WFS1 protein), a transmembrane glycoprotein primarily localized to the endoplasmic reticulum (ER) (22). While WFS1 is expressed ubiquitously, its expression is notably higher in neurons and pancreatic β cells (1, 22). WFS1 protein plays a central role in regulating cellular calcium homeostasis, particularly facilitating Ca^2+^ transfer from the ER to mitochondria through interactions with neuron calcium sensor 1 (NCS1) (23–25). In an in-silico model, it has been demonstrated that the p.Ala684Val variant, present in six of the 11 cases reported here, destabilizes the WFS1 alpha helix, disrupting its interaction with NCS1 (7). This disruption decreases NCS1 levels and leads to downstream mitochondrial respiratory chain dysfunction (23, 24). Additionally, WFS1 interacts with the inositol 1,4,5- triphosphate receptor (IP3R) Ca2+ channel, which is critical for intracellular Ca2+ balance (26). Pathogenic WFS1 variants can impair this interaction, resulting in Ca2+ imbalance and triggering mitophagy, ultimately contributing to cellular degeneration (26). Beyond its role in calcium homeostasis, WFS1 also functions as a negative regulator of the unfolded protein response (UPR) by interacting with activating transcription factor 6α (ATF6α) (27). Thus, loss of function of WFS1 deregulates ER stress pathways, exacerbating cellular ER stress and dysfunction. (27, 28). Notably, the C-terminal domain of WFS1 binds to the ER-localized Na+/K+ ATPase beta-1 subunit (ATP1B1) and facilitates its localization to the cell surface (29–31). Interestingly, four of the five variants in this study (p.Ala684Val, p.Asp797Tyr, p.Gln819Pro, and p.Glu864Lys) are positioned within the ER lumen domain, where they may disrupt this interaction. Studies in mice have shown that a homozygous p.Glu864Lys variant results in defective ATP1B1-WFS1 interactions, leading to impaired endocochlear potential, stria vascularis dysfunction, and neurosensory epithelium abnormalities (31). It is plausible that the other ER lumen variants could similarly affect ATP1B1 function, contributing to sensorineural hearing loss . These molecular insights provide a framework for understanding the clinical manifestations observed in our cohort, particularly the characteristic patterns of optic atrophy and sensorineural hearing loss.

Our findings contribute to a deeper understanding of optic atrophy and sensorineural hearing loss progression in patients with autosomal dominant WFS1-related disorders. The clinical and genetic heterogeneity observed in this cohort highlights the importance of detailed phenotypic and molecular characterization for these patients. Although BCVA remained relatively stable over time, RNFL thickness measurements revealed significant thinning, which correlated more with disease severity than with chronological age. Notably, the p.Gln819Pro variant, reported here for the first time, appears to be associated with less severe manifestations, suggesting potential genotype-phenotype correlations. This study is limited by its retrospective design and the variability in BCVA and RNFL measurements collected from different clinical sites. Nonetheless, these findings provide a foundation for future prospective natural history studies and mechanistic investigations into the pathophysiology of autosomal dominant WFS1-related disorders. Such efforts will be crucial for developing targeted therapeutic strategies and improving clinical management for these patients.

## Author Contributions

FU conceived the investigation design and FU and DH initiated the investigation. JPR, AFT, DH, BC, LO, CB, SH, and FU collected information from patients and databases. JPR, and AFT performed the data analysis. JPR, DH, and FU wrote the manuscript. All authors contributed to the article.

## Data Availability

All data produced in the present work are contained in the manuscript

## Acknowledgments

We sincerely thank the members of the Washington University Wolfram Syndrome and Related Disorders Clinic and Research Team (https://wolframsyndrome.wustl.edu) for their invaluable support. We are especially grateful to all participants in the Wolfram Syndrome and Related Disorders International Registry, Clinical Study, and Clinical Trials for their time, dedication, and commitment to advancing research. We extend special thanks to Dr. Yunshuo Caroline Tang for her expert guidance on neuro-ophthalmological analyses and to Ms. Julie Huecker for her exceptional statistical support.

